# Community social vulnerability and access to medications for opioid use disorder within the continental US: A cross-sectional study

**DOI:** 10.1101/2021.09.30.21264351

**Authors:** Paul J. Joudrey, Marynia Kolak, Qinyun Lin, Susan Paykin, Vidal Anguiano, Emily A. Wang

## Abstract

The COVID-19 pandemic, like past natural disasters, was associated with significant disruptions in medications for opioid use disorder services and increased opioid overdose and mortality. We examined the association between community vulnerability to disasters and pandemics and geographic access to each of the three medications for opioid use disorder within the continental US and if this association was impacted by urban, suburban, or rural classification. We found communities with greater vulnerability did not have greater geographic access to medications for opioid use disorder and the mismatch between vulnerability and medication access was greatest in suburban communities. Rural communities had poor geographic access to all three medications regardless of vulnerability. Future disaster preparedness planning should include anticipation of access to medications for opioid use disorder and better match the location of services to communities with greater vulnerability to prevent inequities in opioid overdose deaths.

## Introduction

US opioid overdose deaths increased within both urban and rural communities and continue to rise in the context of the 2019 novel coronavirus (COVID-19) pandemic.(1,2) Treatment with the three US Food and Drug Administration approved medications for opioid use disorder (MOUD), buprenorphine, methadone, and extended-release naltrexone, can reduce opioid overdose deaths,(3–5) but they are not equivalent or interchangeable.(6) Buprenorphine is a partial opioid agonist available within office-based settings via DATA 2000 waiver.(6,7) Methadone is a full opioid agonist but can only be provisioned at federally certified opioid treatment programs (OTP).(6,7) Extended-release naltrexone is an opioid antagonist and is typically dispensed by a specialty pharmacy and administered by the prescriber.(6,7) Given the differences in pharmacology and delivery, the three MOUD should be accessible in all communities to facilitate treatment individualization and maximization of treatment retention.(6) However, there is a shortage of MOUD services, disproportionately impacting rural communities and contributing to racial inequities in access.(8,9) Most US patients with opioid use disorder (OUD) never initiate a MOUD.(6,10)

Evidence suggests COVID-19 and other recent natural disasters exacerbated the shortage of MOUD services(11–15) and were associated with increased opioid overdose and chronic disease mortality.(2,12,14,16) All-cause mortality increased 62% in Puerto Rico following Hurricane Maria and around one third of excess deaths were attributable to healthcare service disruptions.(16) In the context of COVID-19, US opioid overdose deaths increased 29% from November 2019 to November 2020.(2) Despite efforts to mitigate the impact of COVID-19 on MOUD services, such as increased telemedicine for OAT and relaxation of methadone take home dosing requirements,(15,17) a reduction in locations accepting new methadone patients and long wait times to initiate a medication were observed during the pandemic.(15)

With the ongoing COVID-19 pandemic and the expected increased frequency of climate change related extreme weather events,(18,19) it is important to examine how a community’s ability to respond to natural disasters and infectious disease outbreaks is associated with current access to MOUD, especially given the already uneven access to the medications. The World Health Organization and the US Substance Abuse and Mental Health Services Administration (SAMHSA) have recommended state and local agencies develop disaster plans for patients receiving methadone and buprenorphine.(20–22) But no past studies have examined the relationship between access to each medication and a community’s ability to respond to disaster or infectious disease outbreaks. Identifying communities with greater vulnerability to disasters and pandemics and low access to MOUD could inform interventions and policies aiming to expand MOUD access and mitigate future disparities in mortality. Therefore, we examined the association between social vulnerability and access to each of the three MOUDs within the continental US and if this association was impacted by community urban-rural classification.

## Methods

### Study overview and data sources

We completed a cross-sectional geospatial analysis within the continental US. To measure a community’s ability to respond to natural disasters and infectious disease outbreaks, we obtained 2018 census tract social vulnerability index (SVI) data from Centers for Disease Control and Prevention.(23) The SVI is a validated measure of community vulnerability to natural (e.g. hurricane or infectious disease) or human-caused stressors (e.g. chemical spill).(24) The SVI is derived from 15 US Census Bureau American Community Survey variables and measures overall vulnerability of a census tract and vulnerability across four specific themes: 1) Socioeconomic status (below poverty, unemployed, income, no high school diploma), 2) Household composition and disability (aged 65 or older, aged 17 or younger, older than age 5 with a disability, single-parent households), 3) Minority status and language (minority, speak English “less than well”), and 4) Housing type and transportation (multi-unit structures, mobile homes, crowding, no vehicle, group quarters such as worker dormitories, skilled nursing facilities, or college dorms).(23) The SVI assigns each tract a score based on percentile rank (scored 0 to 1 with 1 representing the highest vulnerability).(23) The SVI was found to predict disaster related property damage and fatalities over a 12-year period among 10 southeastern states and was included within the inter-agency US Climate Resilience Toolkit to facilitate disaster preparedness planning.(25) In the context of COVID-19, increasing SVI scores were associated with increased community COVID-19 cases and deaths and lower rates of COVID-19 vaccination.(26–28)

We measured geographic access to the three MOUD using the following data sources. The primary data source was the SAMHSA Behavioral Health Treatment Services Locator for all substance use treatment clinics providing methadone and extended-release naltrexone (derived from the 2019 National Survey of Substance Abuse Treatment Services) and DATA 2000 waiver buprenorphine providers.(29) Also, because extended-release naltrexone may be provisioned outside of substance use treatment clinics, we obtained location data on all clinicians registered with the pharmaceutical manufacturer as providing extended-release naltrexone from the “Find a treatment provider” website on August 29, 2020.(30)

To provide a comparison of accessing treatment services for another chronic disease necessitating thrice weekly visits, we obtained dialysis center location data from the Centers for Medicare and Medicaid Services’ (CMS) Dialysis Facility Compare database on May 12, 2020.(31)

### Study population

We included all Zip Code Tabulation Areas (ZCTA) within the continental US. Created by the US Census Bureau, ZCTAs are generalized aerial representations of populated US Postal Service ZIP Code service areas.^28^ We used ZCTAs because they are often the smallest geographic unit available to health researchers, in contrast to tract level which may show more neighborhood variation. We excluded Washington, D.C. ZCTAs because the travel cost matrix used for drive time estimation only included US states.

### Dependent Variable

Our primary outcome was drive time in minutes from the population weighted center of the ZCTA to the ZCTA of the nearest treatment location for each treatment type: buprenorphine, methadone, extended-release naltrexone, and dialysis. We excluded treatment locations outside the continental US. There are a limited number of street address locations which cannot be assigned to a ZCTA,(32) and we excluded any treatment location without an assigned ZCTA. To calculate ZCTA access outcomes nationally, we generated an origin-destination matrix of travel times along the street network using Open Source Routing Machine (OSRM) to ZCTAs within 100km.(33) To calculate drive time to the nearest treatment location, we used the *spatial_access* Python package,(33) using the travel time matrix. For origin ZCTAs also containing the treatment destination, the drive time was estimated as 0 minutes.

Secondary outcomes were the total number of treatment locations within or near a ZCTA and number of MOUD types (0 to 3). Treatment locations within a 30-minute drive time of the population weighted center of the ZCTA for each treatment type were tabulated with the *spatial_access* package.(33) We used a 30-minute drive time threshold to represent the number of services available in a ZCTA because 30 minutes is a widely accepted standard for acceptable geographic access for Medicaid beneficiaries and has been used to examine access to methadone for people with OUD and dialysis for people with end stage renal disease.(34–37) To account for differences in the size and age of the population at risk of OUD, we also created a count of treatment locations within a 30-minute drive time per 100,000 adults between ages 18 and 64 for each ZCTA based on the American Community Survey 2018 5-year estimate.

### Independent Variable

Consistent with previous studies,(38) we converted census tract SVI scores into zip code-scale scores using the US Department of Housing and Urban Development U.S. Postal Services ZIP Code Crosswalk.(39) When a zip code spanned multiple census tracts, we calculated weighted average SVI scores using the ratio between the total addresses in each tract over the total addresses in the entire zip code area.(40) We matched all zip codes with SVI scores to their assigned ZCTAs.

### Covariates

Covariates included zip codes classification by urbanicity. We obtained 2010 Rural-Urban Commuting Area (RUCA) codes for zip codes from the US Department of Agriculture and University of Washington Rural Health Research Center.(41) We classified zip codes as either urban (codes 1 and 1.1), suburban (2, 2.1, 4, and 4.1), or rural (all other codes) using RUCA codes. We modified the widely used University of Washington recommendations for RUCA urban-rural classification by first collapsing the large and small rural codes into one category and then identifying codes 3, 5.1, 7.1, 8.1, and 10.1 as rural instead of suburban.(42) This latter change was driven by observations suggesting the traditional University of Washington approach may overestimate urban and suburban areas.(43) We matched all zip codes assigned a RUCA code to their ZCTA. For analyses stratified by urban-rural classification, we excluded ZCTAs without an assigned RUCA code.

### Spatial and Statistical Analysis

First, we identified the count of ZCTAs, total population, total population between ages 18 and 64, and treatment locations within the continental US and the median SVI score among all ZCTAs and within each urban-rural strata. Then we used a Kruskal-Wallis test for comparisons of drive time to the four treatment types and for comparisons by urban-rural strata.

We created correlation matrices among all ZCTAs and among each urban-rural strata, using Spearman’s rank correlation, to examine the relationship between overall SVI and each SVI theme and access to each treatment type. For each correlation matrix, we reversed the direction of the count of treatment locations within a 30-minute drive time so that the direction of correlation for these secondary outcomes were aligned with drive time (positive correlation with SVI meaning greater vulnerability was associated with longer drive time or less treatment locations). We used a Bonferroni correction for multiple comparisons. Given our large sample size, all hypothesis tests were two-sided with an alpha 0.001. We reported correlations of magnitude between –0.09 and 0.09 as no correlation regardless of statistical significance. We completed our analyses in the R software environment (version 4.0.2). University of Chicago Institutional Review Board determined this study did not involve human subjects.

This study has several limitations. First, this study uses the general population between the ages of 18 to 64 to represent treatment need, and this may differ from the location of individuals with OUD. Second, our measures of geographic access to MOUD does not account for the capacity of treatment locations. However, our results are consistent with previous research utilizing a gravity model approach to examine the urban-rural associations between socioeconomic status and access to buprenorphine and methadone.(44) Gravity models aim to account for the capacity of treatment locations and the spatial demand for treatment services. However, gravity model approaches currently require a priori assumptions about the ability to travel for MOUD treatment and the capacity of MOUD treatment locations both of which will vary spatially. Third, our secondary outcomes used a 30-minute travel threshold which likely overestimates availability for people facing transportation barriers. Fourth, our study does not account for mode of transportation (e.g. private vehicle, public bus, etc.) or the impact of traffic or weather. Fifth, because drive time was estimated as zero for ZCTAs containing a treatment location, our results likely underestimate drive time, particularly in urban ZCTAs. However, our secondary outcome is not impacted by this limitation. Sixth, our analysis reflects ZCTA population averages and does not account for variation in social vulnerability within ZCTAs nor does it represent individual-level access.(45,46) Finally, temporal change in vulnerability was beyond the scope of our cross-sectional study but is an important direction for future research.

## Results

Among 32,604 ZCTAs within the continental US, we excluded 170 within Washington, D.C. For analyses stratified by urban-rural classification, we excluded 20 ZCTAs without a RUCA code. Among ZCTAs with a RUCA code (n = 32,584), 10,657 (33%) were urban, 8,067 (25%) were suburban, and 13,860 (43%) were rural (**appendix exhibit 1 and appendix exhibit 2**). Of the over 198 million individuals between age 18 and 64 in ZCTAs with a RUCA code, 76% lived in urban, 15% lived in suburban, and 9% lived in rural ZCTAs. Median overall and theme SVI scores increased (i.e. were more vulnerable) with increasing rural ZCTA classification except for vulnerability due to minority status and language which decreased with increasing rural ZCTA classification (**appendix exhibit 3**).

We matched all buprenorphine (n = 51,191), methadone (n = 1,442), extended-release naltrexone (n = 9,103), and dialysis (n = 7,724) treatment locations to ZCTAs. For analysis by urban-rural classification, we excluded 15 buprenorphine, 2 extended-release naltrexone, and 66 dialysis treatment locations because they resided within the 20 ZCTAs without an assigned RUCA code. Among treatment locations assigned a RUCA code, 3,163 (6%) of buprenorphine, 43 (3%) of methadone, 820 (9%) of extended-release naltrexone, and 656 (8%) of dialysis locations were within rural ZCTAs.

Among all ZCTAs, median drive time to the nearest treatment location was greatest for methadone (35 minutes) and shortest for buprenorphine (16 minutes; *p* <0.001). Only the median drive time to buprenorphine was shorter than median drive time to dialysis (20 minutes). For all treatment types, median drive time increased with increasing rural ZCTA classification (**exhibit 1 and exhibit 2**). Among all ZCTAs, the median count of treatment locations within a 30-minute drive time was greatest for buprenorphine (8) and lowest for methadone (0; *p* <0.001). Among all ZCTAs, the count of methadone and extended-release naltrexone treatment locations within 30 minutes was less than or equal to the count of dialysis centers. For all treatment types, the median count of treatment locations within a 30-minute drive time decreased with increasing rural ZCTA classification. These results were unchanged upon accounting for the number of adults within ZCTAs between the ages 18 and 64. Among all ZCTAs, 152,090,242 (77%) adults ages 18 to 64 lived in a ZCTA with all three MOUD within a 30-minute drive (**appendix exhibit 4**).

### Social vulnerability and treatment access

Among all ZCTAs, greater overall social vulnerability was not associated with greater access (shorter drive times and more available treatment locations) to MOUD. Greater overall social vulnerability was correlated with both longer drive times and less available treatment locations for methadone (correlation 0.10 for drive time and 0.11 for available locations, *p*<0.001). Among all ZCTAs, greater vulnerability due to minority status and language was correlated with greater access to MOUD (correlations between −0.30 and −0.41, *p*<0.001). There was no correlation or a correlation with less access to MOUD for vulnerability due to socioeconomic status (correlations 0.10 to 0.23, *p*<0.001), household composition and disability (correlations 0.25 to 0.39, *p*<0.001), and housing type and transportation (correlations −0.03 to 0.10, *p*<0.001) (**exhibit 3**).

The association between social vulnerability and access to all three types of MOUD varied depending on urban-rural classification. Among rural ZCTAs, increasing overall social vulnerability was correlated with shorter drive times to buprenorphine (correlation −0.10, *p*<0.001) but overall vulnerability was not correlated with other measures of access to MOUD. Vulnerability by theme was not correlated with access among rural ZCTAs, except vulnerability due to socioeconomic status and shorter drive time to buprenorphine (correlation − 0.13, *p*<0.001) and vulnerability due to minority status and language and longer drive time and less available treatment locations for extended-release naltrexone (correlation 0.16 for drive time and 0.15 for available locations, *p*<0.001). While drive time to treatment services largely did not vary by SVI among rural ZCTAs, median drive times were longer in rural ZCTAs as compared with suburban and urban ZCTAs, including among rural ZCTAs in the highest quartile of SVI (**Exhibit 3**).

Among suburban ZCTAs, greater overall vulnerability was correlated (correlations 0.12 to 0.29, *p*<0.001) with both longer drive times and less available locations for all MOUD except drive time to buprenorphine (no correlation). Greater vulnerability due to socioeconomic status, household composition and disability, and housing type and transportation was correlated with both longer drive times and less available locations (correlations 0.13 to 0.30, *p*<0.001), except for drive time to buprenorphine for all themes and drive time to extended-release naltrexone for vulnerability due to housing type and transportation. Among suburban ZCTAs, the median drive time to methadone increased from the lowest quartile of vulnerability to the highest quartile for socioeconomic status (32 to 47 minutes), household composition and disability (33 to 45 minutes), and housing type and transportation (35 to 45 minutes) (**Exhibit 4**).

Among urban ZCTAs, greater overall vulnerability was correlated (correlations −0.29 to −0.10, *p*<0.001) with shorter drive times for all MOUD but was not correlated with available treatment locations. Greater vulnerability due to minority status and language was correlated with shorter drive times and more available locations for MOUD (correlations −0.39 to −0.24, *p*<0.001).

Similar to geographic access to MOUD, among all ZCTAs, overall social vulnerability was not associated with greater access to dialysis for ESRD. Greater vulnerability due to household composition and disability was associated with less access to dialysis (correlations 0.23 for drive time 0.35 for available locations, *p*<0.001), while vulnerability due to minority status and language was associated with greater access (correlations −0.41 for drive time and −0.43 for available locations, *p*<0.001). Upon stratifying ZCTAs by rural-urban status, within rural ZCTAs there was no correlation between overall social vulnerability and access to dialysis. Among suburban ZCTAs, greater overall vulnerability was associated with shorter drive times and but less available dialysis locations (correlation = – 0.10 and 0.12, respectively, *p*<0.001). Among urban ZCTAs, greater overall vulnerability was associated with shorter drive time times and more available treatment locations (correlation = –0.30 for drive time and −0.10 available locations, *p*<0.001).

## Discussion

In this cross-sectional geospatial analysis within the continental US, zip codes with greater social vulnerability did not have greater geographic access to each of the three MOUD, showing the degree to which the US falls short of ensuring equitable access to all MOUD, especially during natural disasters. Consistent with an emerging “opioid treatment desert” literature,(47) nearly one quarter of the continental US population live in zip codes without access to all three MOUD when using a conservative 30 minute travel threshold. We build upon past work by showing that urban-rural inequities were present for all measures of access to buprenorphine and methadone, as well as extended-release naltrexone, including drive time, count of nearby locations, and count of locations per population at risk.(8,44,48) Drive times were significantly longer for methadone and extended-release naltrexone relative to dialysis centers, despite the prevalence of OUD being greater than ESRD.(8,49,50)

A novel finding of this study is that the mismatch between overall social vulnerability and the location of MOUD services was greatest in suburban zip codes as compared to rural and urban zip codes and does not exist for people with end stage renal disease. In examining the specific domains of social vulnerability, living in suburban communities with lower socioeconomic status or in households with more children, seniors, or individuals with disabilities was associated with less geographic access to methadone and extended-release naltrexone. And in contrast, geographic access was largely not associated with social vulnerability in rural zip codes because geographic access to MOUD was uniformly poor. Living in urban zip codes with greater social vulnerability due to higher proportion of residents of racial and ethnic minority status and non-English speakers was associated with greater geographic access to MOUD, suggesting that geographic access may not be as important of a barrier to MOUD treatment in these communities. One important caveat is that geography is just one dimension of access;(51) MOUD access is also impacted by stigma, affordability, accommodation, capacity, and more, and future research should examine the interaction of these factors with geographic access in vulnerable communities.

We improve upon previous research by utilizing two measures of small area (zip code tabulation area) geographic access (drive time and count of near locations) to all three types of MOUD, while incorporating two sources of extended-release naltrexone location data. Our results are consistent with previous research showing communities with lower socioeconomic status have less geographic access to methadone and buprenorphine,(44) and we extend these findings to extended-release naltrexone. By examining the four SVI themes, we find that communities vulnerable due to housing type and transportation also do not have greater geographic access to MOUD and may experience increased mortality during natural disasters and pandemics. While this was also true for vulnerability due to households with children, the elderly and the disabled, its relevance for OUD is less clear because the prevalence of OUD is both lower (i.e. elderly) and higher (i.e. individuals with disabilities preventing entry into the workforce) among subpopulations within this theme.

Our results call into question current disaster preparedness of OUD services and indicate the need to develop proactive measures to increase services within communities with greater vulnerability in the event of a disaster. Expanding OUD services is especially important in vulnerable suburban areas and across rural communities. Methadone should be a priority for innovation given it was the greatest barrier to ensuring access to each MOUD in all communities. Continuing changes in MOUD services during COVID-19, such as telehealth for opioid agonist treatment and increased methadone take home allowances, present opportunities to modify the geographic reach of services.(52) Further, reducing restrictions on medication units (OTP affiliated satellite locations for methadone administration and dispensing) and the recent end to the moratorium on new mobile methadone vans may also mitigate urban-rural inequalities and increase MOUD services within vulnerable communities if strategically implemented.(34,53) Lastly, allowing methadone treatment outside of OTPs could greatly expand access, but office-based methadone would require federal and state regulatory changes for wide spread adoption.(54,55) Currently, SAMHSA, the Drug Enforcement Agency, and State Opioid Treatment Authorities disaster planning for methadone treatment prioritize coordination among existing OTPs.(56) But coordination among OTPs alone in event of a disaster has been insufficient to ensure access to methadone,(15) especially in communities without a nearby OTP or alternate OTP. Canada increased the flexibility of its federal and provincial methadone regulations, including allowing methadone in office-based settings and medication dispensation within pharmacies, resulting in an expansion of methadone treatment services.(57) More flexible methadone regulation at the federal and state level within the US is likely required if the identified inequities in disaster preparedness are to be mitigated.

In conclusion, zip codes within the continental US with greater social vulnerability did not have greater geographic access to buprenorphine, methadone, or extended-release naltrexone. The mismatch between social vulnerability and the location of MOUD services was greatest in suburban zip codes, but rural zip codes had longer drive times to all three MOUD regardless of vulnerability. MOUD policy and delivery innovations need to address urban-rural inequities and better match the location of services to communities with greater social vulnerability to prevent inequities in opioid overdose deaths during future natural disasters.

## Data Availability

All data is available upon request.

## ACKNOWLEDGMENTS

We would like to thank Moksha Menghaney for assistant managing the SAMHSA data and Nathan Kim for his assistant obtaining the data on extended-release naltrexone.

---

**Exhibit 1.** Measure of Access to Medications for Opioid Use Disorder among ZCTAs within the Continental US.

| Variable | N | Overall, N = 32,584<sup>1</sup> | Rural, N = 13,860<sup>1</sup> | Suburban, N = 8,067<sup>1</sup> | Urban, N = 10,657<sup>1</sup> | P-value<sup>2</sup> |
| --- | --- | --- | --- | --- | --- | --- |
| **Time (Methadone)** | 29,379 | 35.00 (16.00, 60.00) | 62.00 (44.00, 84.00) | 39.00 (27.00, 54.00) | 13.00 (7.00, 20.00) | <0.001 |
| **Time (Naltrexone)** | 32,143 | 22.00 (9.00, 40.00) | 38.00 (23.00, 59.00) | 25.00 (16.00, 37.00) | 7.00 (0.00, 12.00) | <0.001 |
| **Time (Dialysis)** | 32,456 | 20.00 (8.00, 34.00) | 33.00 (22.00, 49.00) | 22.00 (15.00, 32.00) | 6.00 (0.00, 11.00) | <0.001 |
| **Time (Buprenorphine)** | 32,557 | 16.00 (0.00, 30.00) | 28.00 (17.00, 43.00) | 19.00 (10.00, 28.00) | 0.00 (0.00, 7.00) | <0.001 |
| **count (Methadone)** | 32,584 | 0.00 (0.00, 2.00) | 0.00 (0.00, 0.00) | 0.00 (0.00, 1.00) | 3.00 (1.00, 8.00) | <0.001 |
| **count (Naltrexone)** | 32,584 | 2.00 (0.00, 10.00) | 0.00 (0.00, 1.00) | 1.00 (0.00, 4.00) | 20.00 (8.00, 42.00) | <0.001 |
| **count (Dialysis)** | 32,584 | 2.00 (0.00, 8.00) | 0.00 (0.00, 1.00) | 2.00 (0.00, 3.00) | 16.00 (7.00, 36.00) | <0.001 |
| **count (Buprenorphine)** | 32,558 | 8.00 (1.00, 53.00) | 1.00 (0.00, 5.00) | 6.00 (1.00, 19.00) | 117.00 (43.00, 269.00) | <0.001 |
| **count (Methadone) per 100K** | 32,360 | 0.00 (0.00, 30.81) | 0.00 (0.00, 0.00) | 0.00 (0.00, 20.29) | 31.65 (10.88, 99.26) | <0.001 |
| **count (Naltrexone) per 100K** | 32,424 | 61.51 (0.00, 302.00) | 0.00 (0.00, 117.92) | 43.73 (0.00, 272.48) | 201.56 (72.35, 544.08) | <0.001 |
| **count (Dialysis) per 100K** | 32,442 | 69.41 (0.00, 253.49) | 0.00 (0.00, 127.71) | 61.96 (0.00, 241.18) | 162.71 (66.93, 427.03) | <0.001 |
| **count (Buprenorphine) per 100K** | 32,444 | 378.79 (24.27, 1,619.43) | 78.22 (0.00, 587.15) | 286.74 (31.99, 1,304.27) | 1,211.60 (441.48, 3,468.37) | <0.001 |

<sup>1</sup>Median (IQR). <sup>2</sup>Kruskal-Wallis rank sum test.
Source: The primary data source was the SAMHSA Behavioral Health Treatment Services Locator for all substance use treatment clinics providing methadone and extended-release naltrexone (derived from the 2019 National Survey of Substance Abuse Treatment Services) and DATA 2000 waiver buprenorphine providers.(29) We also obtained location data on all clinicians registered with the pharmaceutical manufacturer as providing extended-release naltrexone from the “Find a treatment provider” website on August 29, 2020.(30)

**Exhibit 2.**
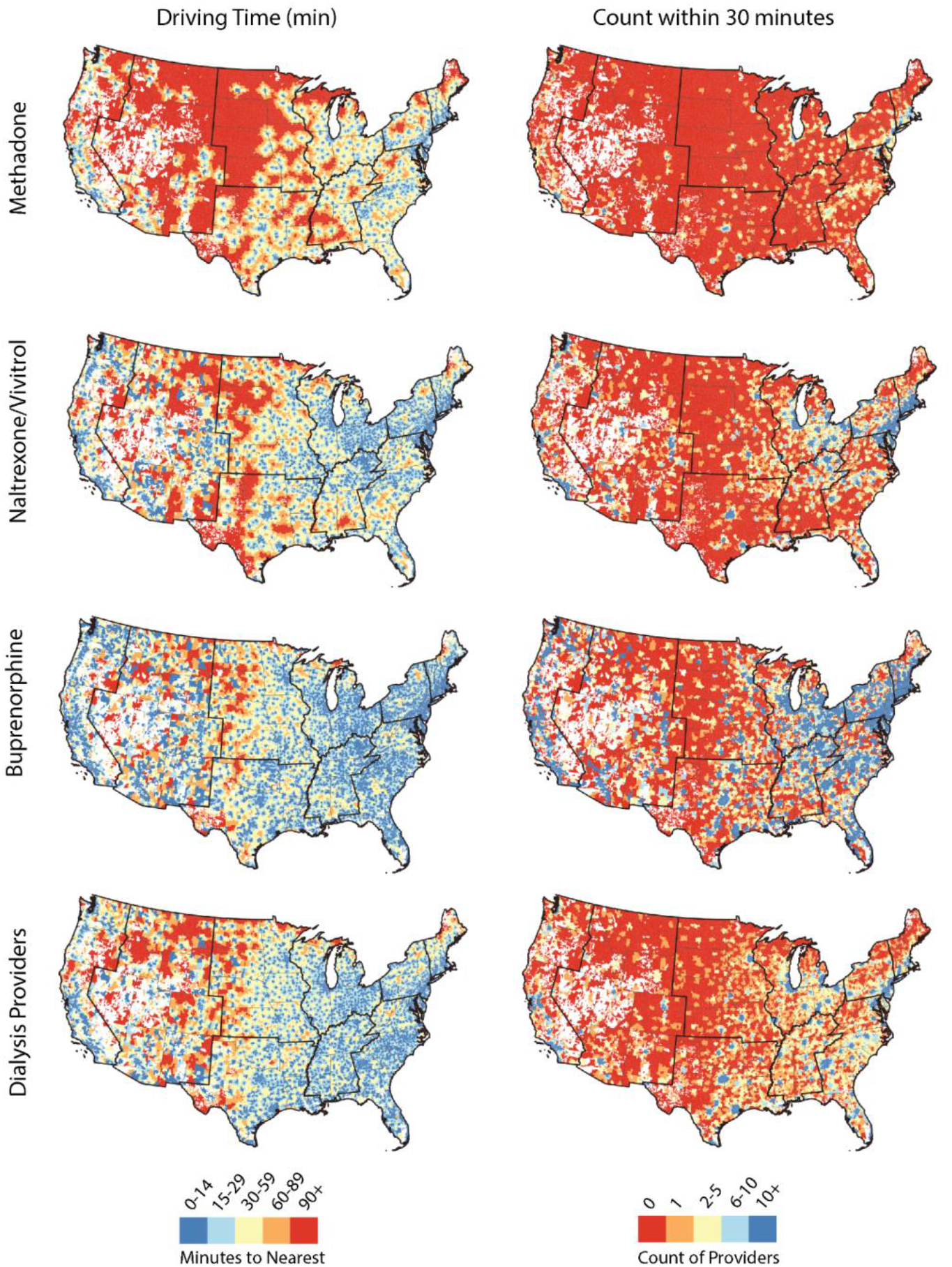
Driving time to the nearest MOUD resource (buprenorphine, extended-release naltrexone, methadone, and dialysis center) and count of resources within 30 minutes driving time in the continental US in 2020 with Census Regions bolded. Driving time map breaks were derived based on using 90 minutes as a conservative benchmark for the highest threshold (lowest level of access) for reaching MOUD providers. Based on existing access literature, map breaks were chosen to show variation of accessibility within 15 minutes, 30 minutes (benchmark of accessibility in existing dialysis literature), 60 minutes, 90 minutes, and greater than 90 minutes.

**Exhibit 3.**
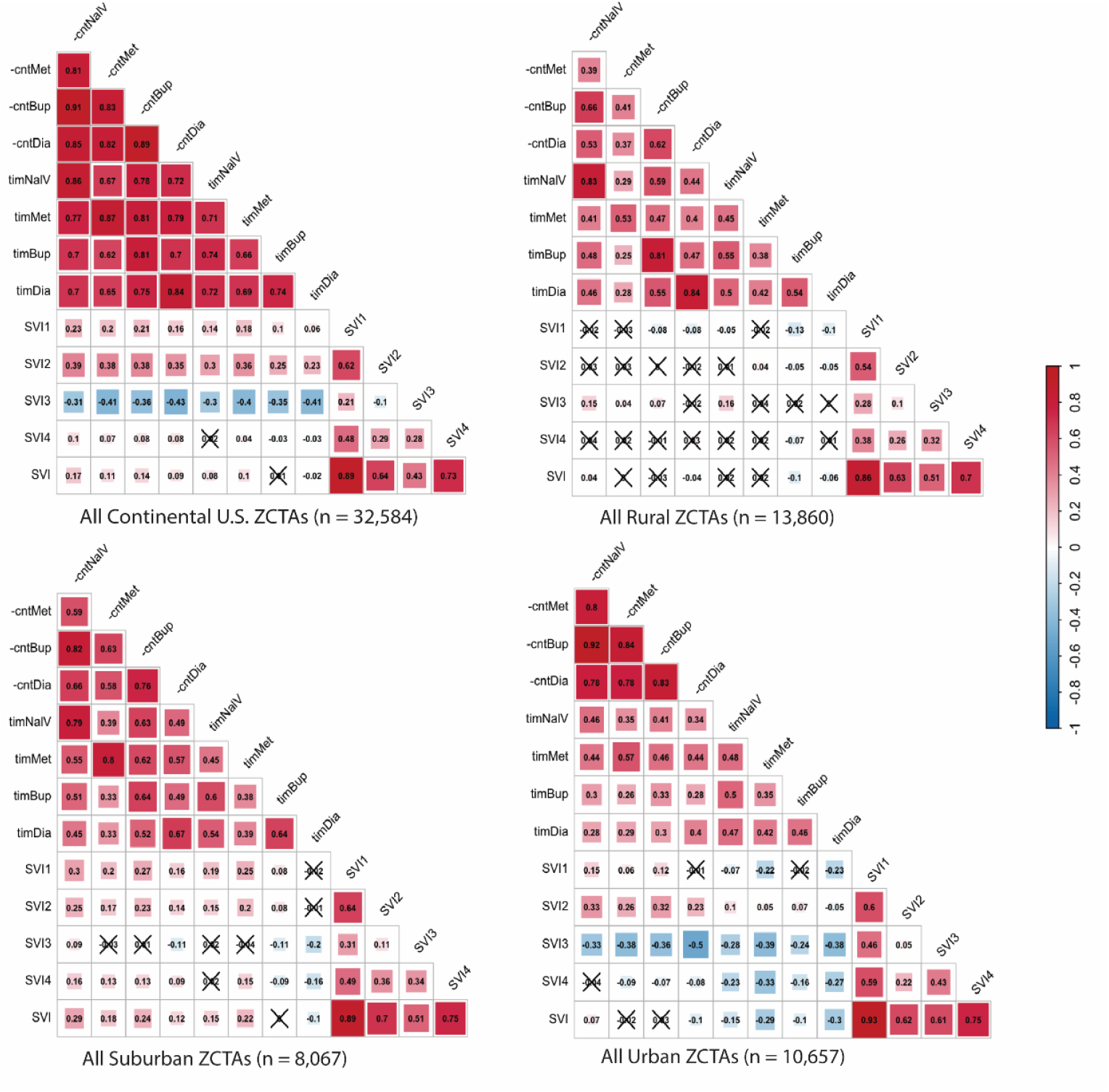
Correlation between social vulnerability and access to medications for opioid use disorder and dialysis centers in the continental US in 2020. The shading depends on the magnitude of correlation coefficient, with red indicating positive correlations and blue indicating negative correlations.

**Exhibit 4.** Summary Statistics of Driving Time Access Metrics across SVI Themes.

| Median Drive Time to the Nearest Methadone, Buprenorphine, and Naltrexone |  |  |  |  |  |  |  |  |  |  |  |  |  |
| --- | --- | --- | --- | --- | --- | --- | --- | --- | --- | --- | --- | --- | --- |
| SVI Theme |  | Rural |  |  |  | Suburban |  |  |  | Urban |  |  |  |
| *Increasing vulnerability* | Quar. | Met | Nal | Dial | Bup | Met | Nal | Dial | Bup | Met | Nal | Dial | Bup |
| **Socioeconomic Vulnerability**<br>(SVI Theme 1) | 1 | 60 | 40 | 35 | 31 | 32 | 22 | 22 | 18 | 15 | 8 | 8 | 0 |
|  | 2 | 65 | 39 | 35 | 29 | 36 | 23 | 23 | 19 | 13 | 7 | 6 | 0 |
|  | 3 | 62 | 36 | 32 | 26 | 42 | 25 | 21 | 19 | 12 | 6 | 5 | 0 |
|  | 4 | 61 | 38 | 31 | 25 | 47 | 32 | 23 | 22 | 9 | 6 | 4 | 0 |
| **Household Composition & Disability**<br>(SVI Theme 2) | 1 | 57 | 36 | 33 | 27 | 33 | 21 | 22 | 17 | 12 | 6 | 7 | 0 |
|  | 2 | 62 | 39 | 34 | 29 | 37 | 24 | 23 | 19 | 13 | 7 | 6 | 0 |
|  | 3 | 63 | 37 | 33 | 28 | 40 | 26 | 22 | 20 | 13 | 8 | 6 | 0 |
|  | 4 | 63 | 39 | 32 | 27 | 45 | 29 | 23 | 21 | 12 | 8 | 6 | 0 |
| **Minority Status & Language**<br>(SVI Theme 3) | 1 | 61 | 34 | 33 | 27 | 40 | 25 | 24 | 21 | 21 | 14 | 14 | 9 |
|  | 2 | 61 | 37 | 32 | 27 | 38 | 24 | 23 | 19 | 18 | 11 | 11 | 3 |
|  | 3 | 63 | 43 | 32 | 27 | 38 | 26 | 20 | 18 | 14 | 7 | 7 | 0 |
|  | 4 | 64 | 49 | 35 | 30 | 38 | 28 | 19 | 17 | 10 | 5 | 4 | 0 |
| **Housing Type & Transportation**<br>(SVI Theme 4) | 1 | 61 | 36 | 32 | 28 | 35 | 24 | 24 | 20 | 17 | 10 | 9 | 0 |
|  | 2 | 61 | 37 | 32 | 28 | 38 | 25 | 23 | 20 | 14 | 7 | 7 | 0 |
|  | 3 | 62 | 39 | 33 | 28 | 40 | 25 | 21 | 19 | 11 | 5 | 4 | 0 |
|  | 4 | 63 | 39 | 34 | 26 | 45 | 26 | 19 | 16 | 8 | 4 | 4 | 0 |

Source: We obtained 2018 Census tract social vulnerability index (SVI) data from Centers for Disease Control and Prevention.

**Appendix Exhibit 1.**

|  | Total | Urban | Suburban | Rural |
| --- | --- | --- | --- | --- |
| Naltrexone | 9,103 | 7,004 | 1,277 | 820 |
| Methadone | 1,442 | 1,254 | 145 | 43 |
| Buprenorphine | 51,191 | 42,784 | 5,229 | 3,163 |
| Dialysis | 7,724 | 5,828 | 1,174 | 656 |
| ZCTA* | 32,584 | 10,657 | 8,067 | 13,860 |
| Population | 320,026,981 | 240,888,576 | 47,938,471 | 31,199,934 |
| Population 18–64 | 198,264,579 | 151,041,785 | 28,976,347 | 18,246,447 |

\* All ZCTAs were used for access metric calculation, however, not all ZCTAs are included in the USDA RUCA dataset. Specifically, 20 ZCTAs are missing from the USDA zip code RUCA codes. 0.119% of MOUD and dialysis resources are located in these 20 ZCTAs, thus they were not assigned to any RUCA classification. These include 2 out of 9,103 Naltrexone locations, 15 out of 51,191 Buprenorphine locations and 66 out of 7724 Dialysis locations.

**Appendix Exhibit 2.**
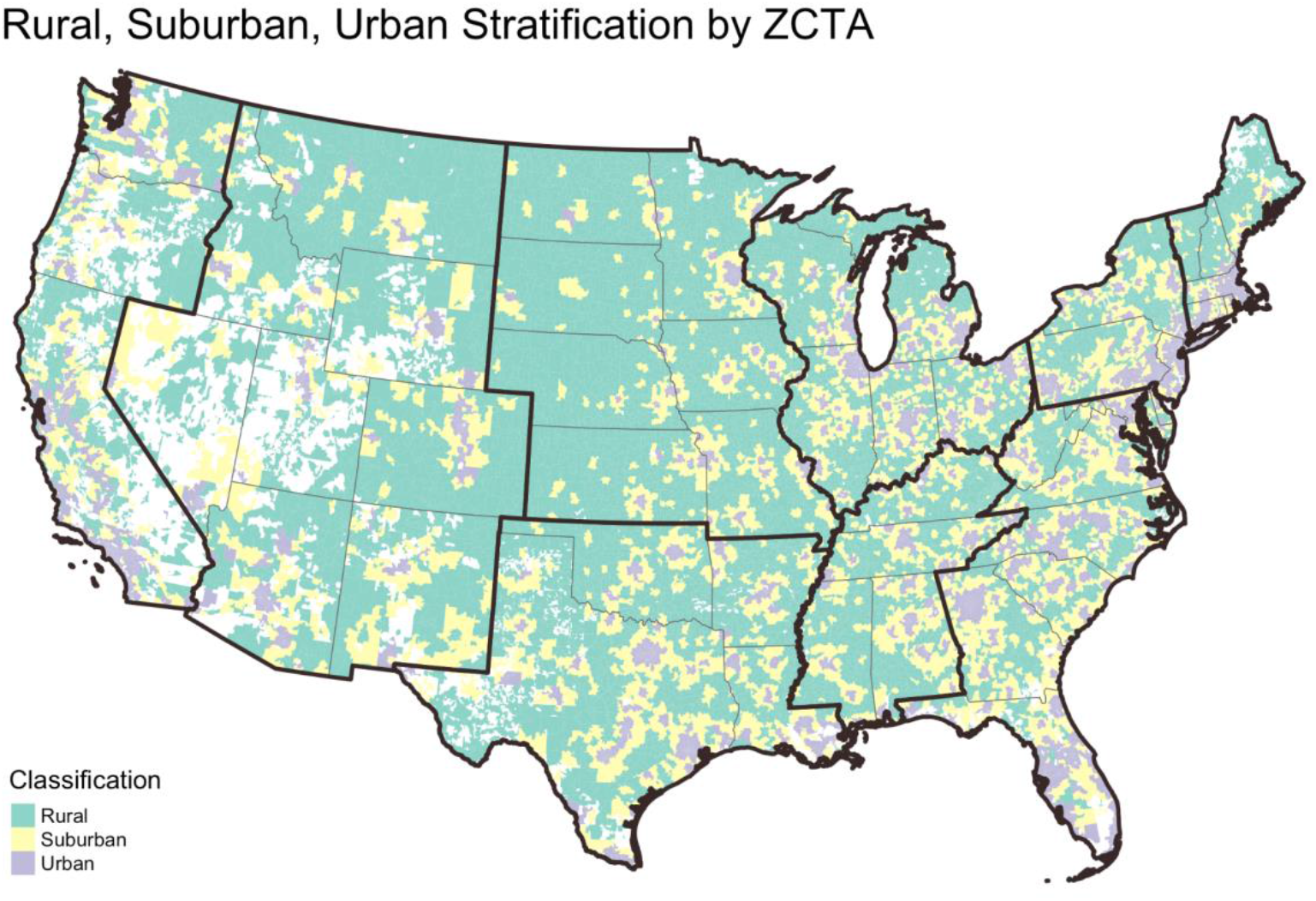
All assigned ZIP Code Tract Areas (ZCTA) in the continental US classified as rural, suburban, or urban, developed based on the U.S. Office of Management and Budget (OMB) RUCA Codes.

**Appendix Exhibit 3.**
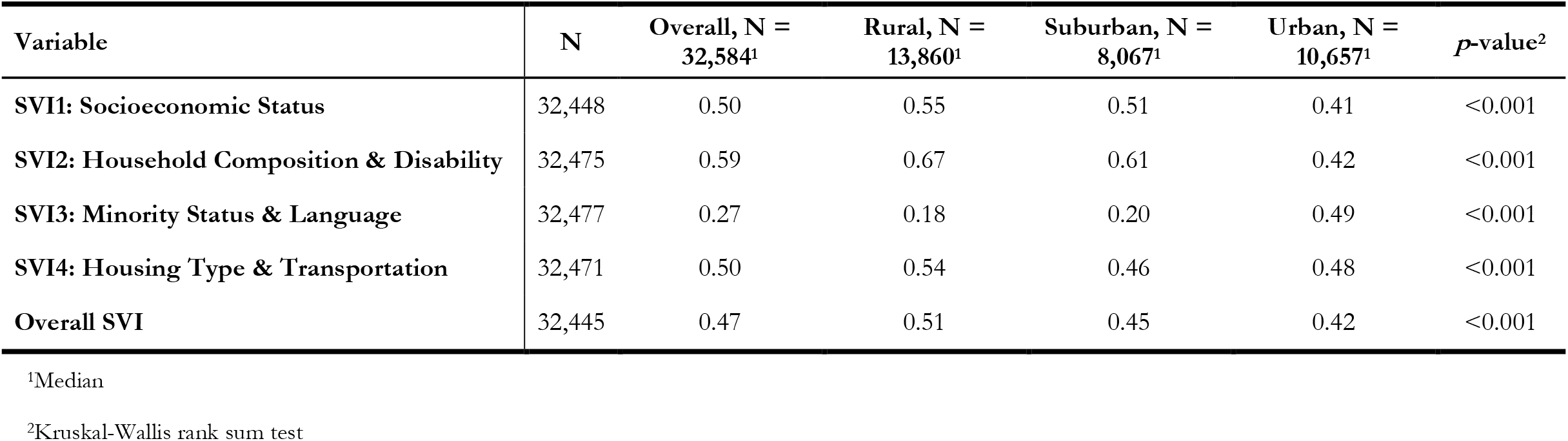

**Appendix Exhibit 4.**
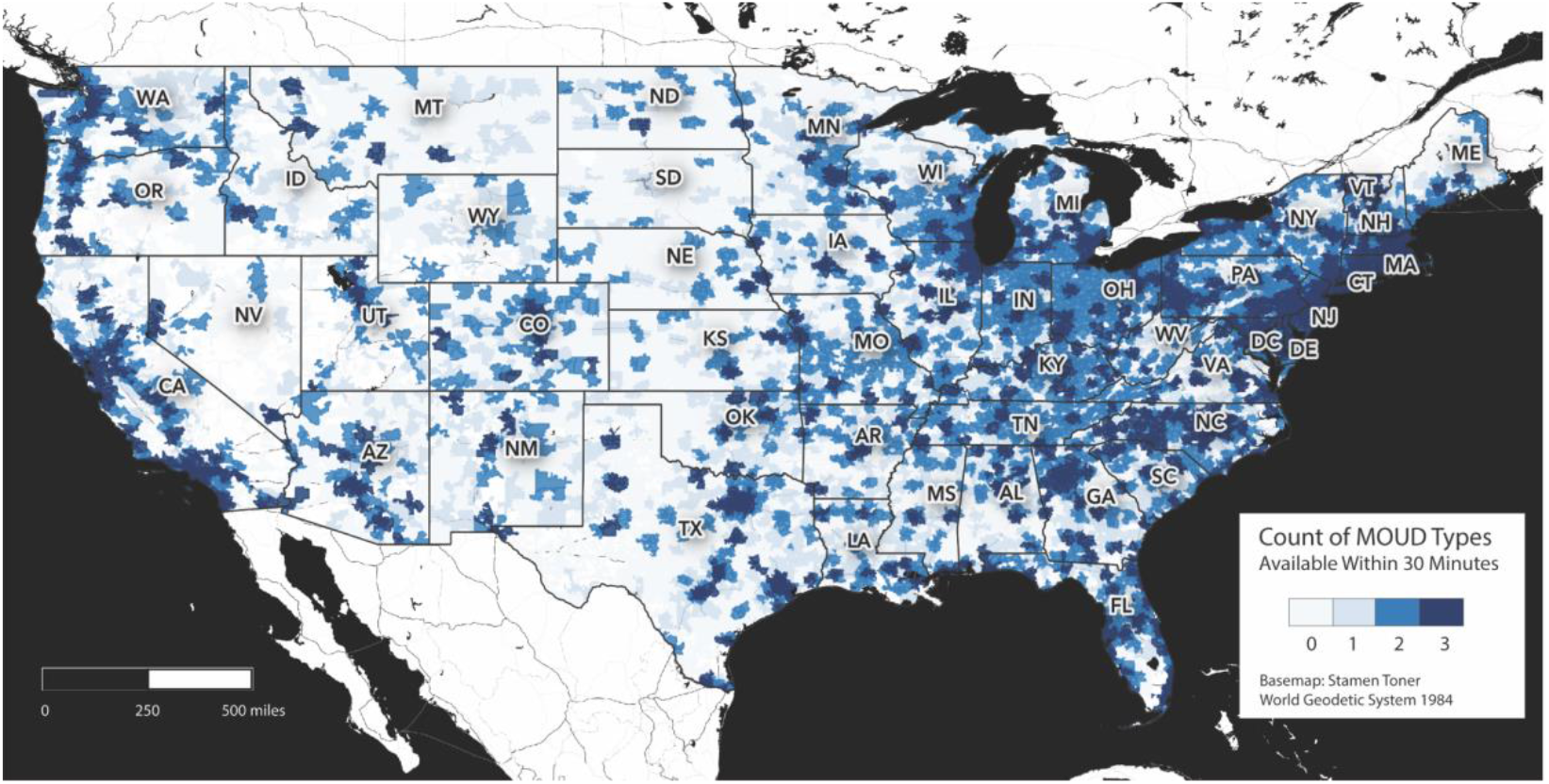
Count of MOUD types within 30 minutes driving time of every population-weighted ZCTA centroid in the continental US.

## Notes

### Competing Interest Statement

The authors have declared no competing interest.

### Funding Statement

Funding for this publication was provided by grant number 5K12DA033312 (P.J.J.), L30 DA052056 (P.J.J.), from the National Institute on Drug Abuse, a component of the National Institutes of Health.
The funding organization had no role in the design and conduct of the study; collection, management, analysis, and interpretation of the data; preparation, review, or approval of the manuscript; and decision to submit the manuscript for publication.

### Author Declarations

Yale University IRB determined this study was not human subjects research.

